# Piloting short Empathetic Refutational Interview modules in clinical training: Two UK studies

**DOI:** 10.1101/2025.08.05.25333036

**Authors:** Dawn Holford, Emma C. Anderson, Harriet Fisher, Virginia C. Gould, Frederike Taubert, Linda C. Karlsson, Stephan Lewandowsky

## Abstract

**Background:** Effective communication from trusted healthcare professionals (HCPs) can increase patients’ acceptance of vaccination. However, many HCPs find these conversations challenging and lack communication confidence when patients express vaccine concerns. The Empathetic Refutational Interview (ERI) is a framework for improving vaccine conversations and addressing vaccine misconceptions. Our objective was to pilot ERI training as continuing medical education to improve HCPs’ vaccine communication confidence.

**Method:** We piloted a short-form (60-90 mins) ERI training module for HCPs in two different UK clinical training settings. In Study 1, the ERI module was run within four immunisation training days and compared to a control communication module of the same length. In Study 2, the ERI module was run as ten stand-alone sessions with no control group. We conducted a mixed-methods evaluation of training impact on participants’ confidence and preparedness for vaccine conversations. We collected self-report measures and qualitative feedback from participants immediately before and after training, and subsequently one and three months post training. We also conducted structured observations of ERI training.

**Results:** We recruited participants from the HCPs (predominantly nurses) who attended the training (Study 1: *n* = 61; Study 2: *n* = 98). Participants showed significant improvements in self-reported communication confidence and preparedness for vaccine conversations after all training modules. Control group participants described improved knowledge of information sources as supporting their confidence, while ERI group participants described improved communication skills and techniques. Participants reported that the ERI provided a helpful framework to structure and practise conversations. Participants and observers felt that more practice time would enhance training.

**Conclusions:** Short training modules can improve HCPs’ confidence in vaccine communication. Having an evidence-based communication structure can help HCPs gain awareness of skills for effective communication about vaccination, not just knowledge around signposting patients to information.

## Introduction

Although vaccination is the safest and most effective way to protect against many diseases^1^, vaccine hesitancy—the refusal or delay of available vaccinations^2^—threatens the success of global immunisation programmes. Declining vaccine coverage has led to outbreaks of vaccine-preventable diseases, e.g., measles outbreaks worldwide^3^.

Healthcare professionals (HCPs) are trusted influencers whose recommendations can encourage vaccination uptake among hesitant patients^4–6^. Communicating effectively about vaccination can require HCPs to address misconceptions and false beliefs^7,8^ while practising an empathetic and participatory engagement style^9,10^. However, it can be challenging for HCPs to communicate in this style^11^. HCPs often receive limited training in vaccine communication, with most educational interventions assuming that hesitancy is due to a patient’s information deficits^10^. This gap in communication skills can affect HCPs’ confidence to recommend vaccination^12^. HCPs may even avoid difficult vaccine conversations if they suspect it might lead to conflict with their patient^11,13^. It is thus important to help HCPs build their skills and confidence to deploy evidence-based best practice to communicate with vaccine-hesitant patients.

### The Empathetic Refutational Interview

One communication intervention that showed promising results in addressing vaccine hesitancy is Motivational Interviewing (MI): MI-trained HCPs have successfully improved the vaccine readiness of patients they speak with^14,15^. However, MI does not provide specific guidance on dealing with misinformation, which HCPs would welcome specific training to address^11,16^. Our study therefore used a communication framework designed to combine the strengths of MI with evidence-based psychological science strategies for refuting misconceptions: the Empathetic Refutational Interview (ERI)^17^.

The ERI is a four-step framework to guide conversations with vaccine-hesitant individuals (see Figure 1). One of its foundations is attitude root theory, i.e., that underlying psychological attributes (attitude roots) motivate specific concerns that individuals may express^18^. The ERI was found to gain more receptivity from vaccine-hesitant individuals compared to an information-giving approach^17^ and HCPs also perceived the ERI as a better way to handle conversations with hesitant patients^19^. ERI-trained HCPs have increased patients’ willingness to vaccinate and vaccine appointments booked after consultations compared to untrained HCPs^20^. However, it is not yet known how best to train HCPs to use the ERI approach.

**Figure 1.**
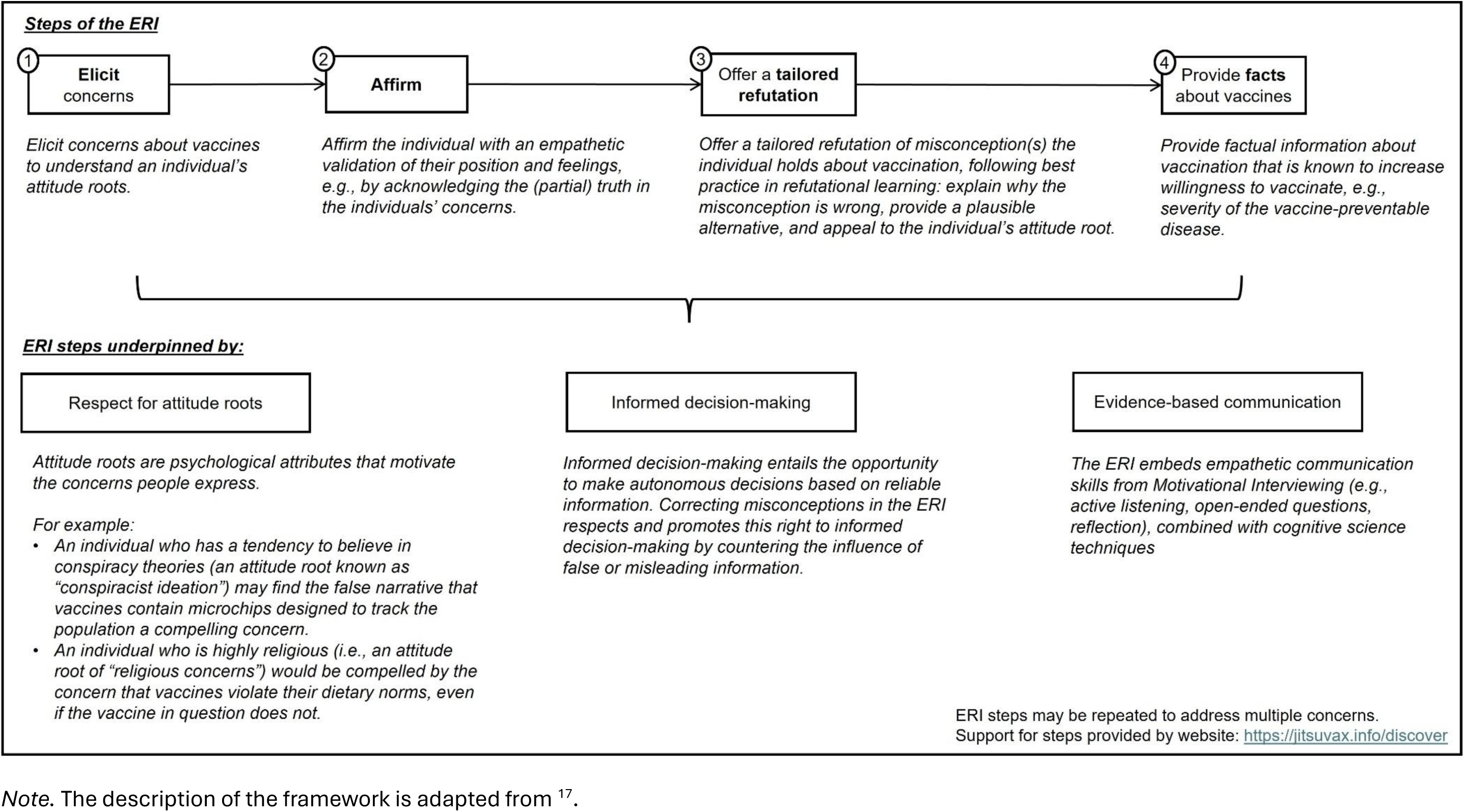
The four-step framework of the Empathetic Refutational Interview (ERI).

### Objective and overview of research

Our objective was to understand the impact of a brief ERI training module on HCPs’ vaccine communication confidence and preparedness to address vaccine misconceptions that patients might bring up. We also assessed HCPs’ qualitative feedback on the training.

We conducted two mixed-methods studies. In Study 1, we delivered the ERI module at the end of an immunisation training day and compared it to an existing communication module of the same length (control group). In Study 2, we delivered the ERI module as a stand-alone session with no control group. We evaluated changes in trainees’ self-reported confidence and skills in each study. We also explored what elements in the modules support HCPs’ learning. Across studies, we collected qualitative data for process evaluation, including questionnaire feedback on training experience and structured observations of the ERI training.

## Study 1

### Methods

#### Study context

We embedded the study within an in-person child immunisation training day run by a regional NHS health organisation in the South West of England. The full day covered theoretical and practical aspects of childhood immunisation and concluded with a 60-minute module on communicating with patients and caregivers. Trainees (Table 1) were randomised to receive this (control) communication module or the ERI module. Data collection took place before and after the communication module. The study involved four separate training days between March and November 2023. Primary quantitative analyses were pre-registered on the Open Science Framework prior to data collection (https://osf.io/7a3rq?view_only=5cddcd0379e445beba0333dbdc4b16c0).

**Table 1.**
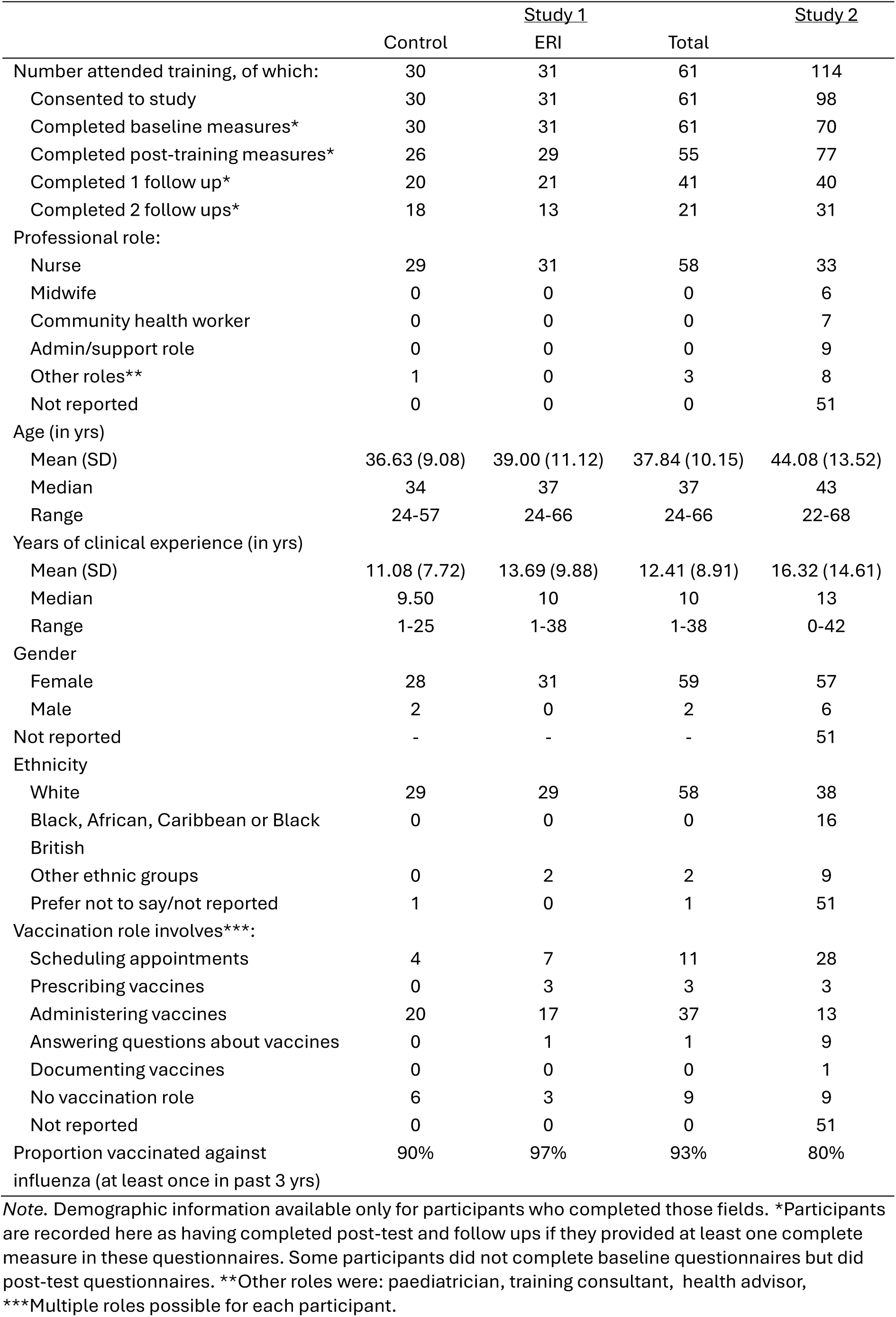
Breakdown of participants in each study and their characteristics.

### Design and procedure

#### Recruitment

Trainees were emailed information about the study before the training day. Participants expressed consent before completing a baseline questionnaire before proceeding to the training day.

Participants attended the full training day all together and were randomly assigned to either the control or the ERI group for the final module, which the groups received in different rooms^1^. After this module, participants completed a post-training questionnaire.

Participants were sent follow-up online questionnaires via an emailed link at one month and three months post-training. Participants were offered the chance to win a £50 shopping voucher for completing each follow-up questionnaire. Participants received three email reminders to complete each follow-up questionnaire, sent at one-week intervals.

### Training modules

#### ERI module

The 60-minute ERI module consisted of a short presentation on the ERI framework, a demonstration video, an introduction to a supporting web resource (https://jitsuvax.info), and two scenario-based exercises where trainees played the role of a HCP or patient to practise using the ERI in conversation, followed by a facilitated discussion. Two members of the research team (EA, HF) delivered the module.

#### Control module

This was an existing 60-minute communication module for the immunisation training day. It consisted of a scenario-based groupwork exercise where trainees discussed potential patient (or carer) concerns about a vaccination in small groups and brought these to a whole group facilitated discussion. The immunisation training co-ordinator delivered this module.

#### Outcome measures

Our primary outcome measures were vaccine communication confidence (three items) and perceived preparedness to refute vaccine misconceptions (six items). Both were measured at baseline, post-training, and one and three-month follow-ups. Both measures used a five-point response scale with higher values indicating greater confidence and preparedness. For each measure, we computed a mean score for each participant. Full wording and descriptive statistics for these measures are provided in Supplementary Material.

#### Vaccine communication confidence

We used three items from the “proactive self-efficacy” section of the International Professionals Vaccine Confidence and Behaviours questionnaire (I-Pro-VC-Be^12^), which ask about HCPs’ commitment and preparedness to discuss vaccines with hesitant patients.

#### Refutation preparedness for vaccine misconceptions

We selected six commonly encountered anti-vaccination arguments^21^. Participants rated how prepared they felt to respond to each argument if a patient brought it up.

#### Secondary outcome measures

Participants also reported the number of influenza vaccinations they received in the last three years, self-reported understanding of ERI techniques, and frequency of use of ERI techniques. We report results for the secondary outcomes in Supplementary Material.

## Results

### Participants

All trainees who attended the training day consented to participate (*n* = 61; participant demographics in Table 1).

### Quantitative analyses

We ran pre-registered between-within analyses of variance (ANOVAs) on each primary outcome measure (Figure 2), comparing study conditions (ERI vs. control; between-subjects) and timing (baseline vs. post-training; within-subjects). We then ran exploratory pairwise comparisons including the two follow-up time points.

**Figure 2.**
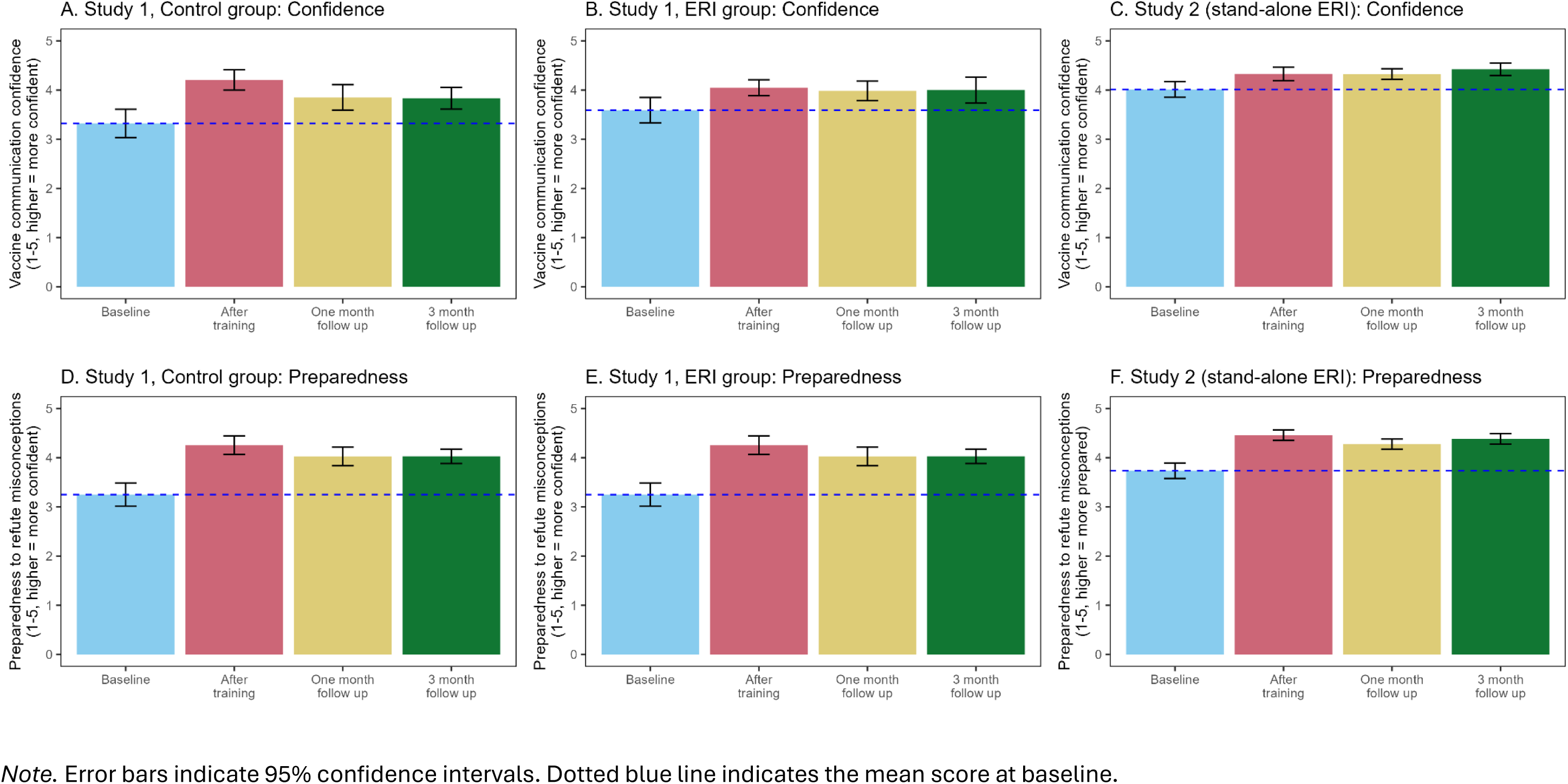
Confidence (top row) and preparedness (bottom row) at baseline and after training (immediately and at follow-ups) in Study 1 and 2.

All participants’ communication confidence improved significantly immediately post-training, *F*(1, 52) = 45.68, *p* < .001, η^2^_P_ = 0.47 (*d* = 0.90). This improvement was not significantly different between conditions, *F*(1, 52) = 2.84, *p* = .098, η^2^_P_ = 0.05 (interaction effect). For participants who completed the follow-ups, pairwise *t*-tests showed that confidence remained significantly higher than baseline at follow-up 1, *t*(40) = 4.22, *p* < .001, *d* = 0.66, and follow-up 2, *t*(30) = 2.88, *p* = .020, *d* = 0.52. Change in confidence did not interact with condition, *F*(3, 60) = 2.49, *p* = .070, η^2^_P_ = 0.11.

All participants’ refutation preparedness increased significantly immediately post-training, *F*(1, 52) = 90.87, *p* < .001, η^2^_P_ = 0.64 (*d* = 1.31). This improvement was also not significantly different between conditions, *F*(1, 52) = 0.18, *p* = .676, η^2^_P_ < 0.01 (interaction effect). For participants who completed the follow-ups, pairwise *t*-tests showed that preparedness remained significantly higher than pre-test at follow-up 1, *t*(39) = 8.34, *p* < .001, *d* = 1.32, and follow-up 2, *t*(30) = 5.87, *p* < .001, *d* = 1.05. Change in refutation preparedness did not interact with condition, *F*(3, 60) = 1.68, *p* = .179, η^2^_P_ = 0.08.

## Study 2

### Methods

#### Study context

We organised ten ERI training sessions between June 2023 and May 2024 in London, the Midlands, and North West UK regions, each lasting 90 minutes. The module took approximately 60 minutes (same as in Study 1), with 15 minutes before and after for primary data collection. Health organisations and local authorities in each region advertised the sessions to HCPs in their catchment areas. Research team members (DH, HF) conducted the sessions in-person except for one delivered online at the request of the partnering health organisation.

#### Design and procedure

All participants attended the ERI module, because no partner organisation could provide a comparative communication module—underscoring the training provision gap. Data collection procedures were similar to Study 1.

## Results

### Participants

Ninety-eight trainees (out of 114) consented to inclusion in the study (see Table 1 for participant demographics)^2^.

### Quantitative analyses

Analyses for this study focus on changes in the primary outcomes of vaccine communication confidence and preparedness to refute vaccine misconceptions (see Figure 2). They were not pre-registered. We used paired samples *t*-tests to analyse baseline and post-training differences, followed by one-way within-subjects ANOVAs testing for changes over the follow-up period.

Compared to baseline, participants’ communication confidence increased significantly post-training, *t*(57) = 2.80, *p* = .007, *d* = 0.37. The ANOVA across the follow-ups was not significant (*p* = .224).

Refutation preparedness increased significantly from baseline to post-training, *t*(57) = 7.42, *p* < .001, *d* = 0.97. The ANOVA across the follow-ups showed a significant overall change in preparedness, *F*(3, 36) = 13.46, *p* < .001, η^2^_P_ = 0.53. Pairwise comparisons showed that preparedness remained significantly higher at follow-up 1 and 2 compared to baseline, respectively: *t(*25) = 3.68, *p* = .004, *d* = 0.72; *t*(20) = 5.82, *p* < .001, *d* = 1.27.

## Process evaluation

### Method

Our process evaluation across both studies consisted of: feedback questionnaires collected post-training and at follow-up, and structured observations of the ERI modules. The evaluation question and observation schedule are provided in Supplementary Material.

#### Feedback questionnaires

Participants were asked to give a quantitative rating of the informativeness, clarity, and usefulness of the session they attended immediately after training. This questionnaire invited open-ended feedback on: usefulness of training content, improvements in understanding, intended use in clinical practice, suggestions for workshop improvement and general feedback. At follow up 1 and 2, participants reported what training elements they had used in clinical practice and what they remembered from training. Data were analysed using a pragmatic qualitative content analysis^22^. A spreadsheet of free-text data was coded for response-types independently by two researchers (DH, EA), and final categories were confirmed via discussion. We used the tm and wordcloud packages in R version 4.3.1^23,24^ to generate word clouds as visual representations of the full set of free-text responses.

#### Structured observations

A member of the research team completed an observation schedule for each ERI session to assess the fidelity of module delivery.

## Results

We report here a content analysis of the free-text responses obtained across studies, highlighting differences between control and ERI participants. We then provide brief results of the structured observations. Broad categories of responses are reported in the text, with illustrative quotations presented in the respective tables (Table 2).

**Table 2.**
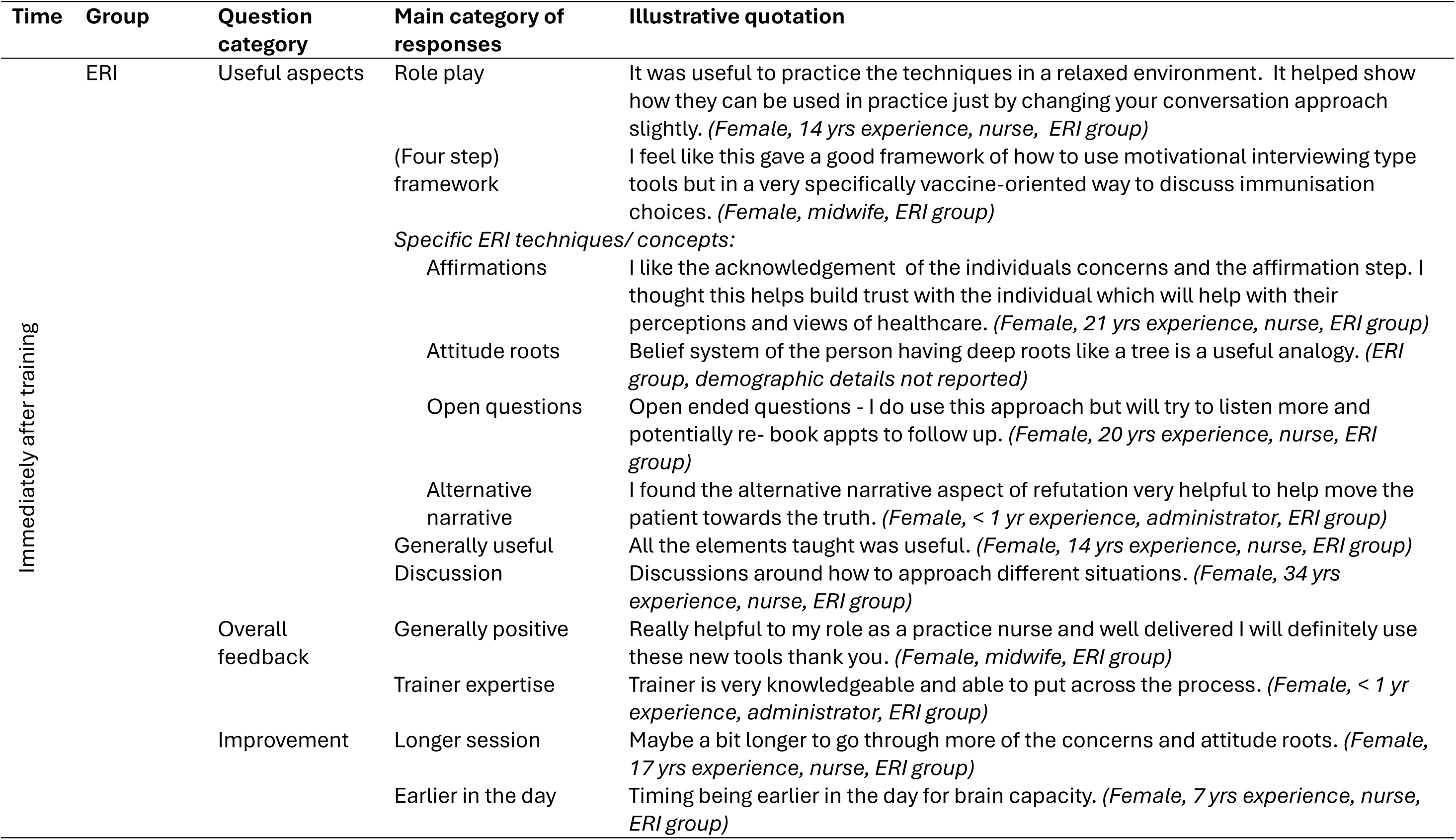
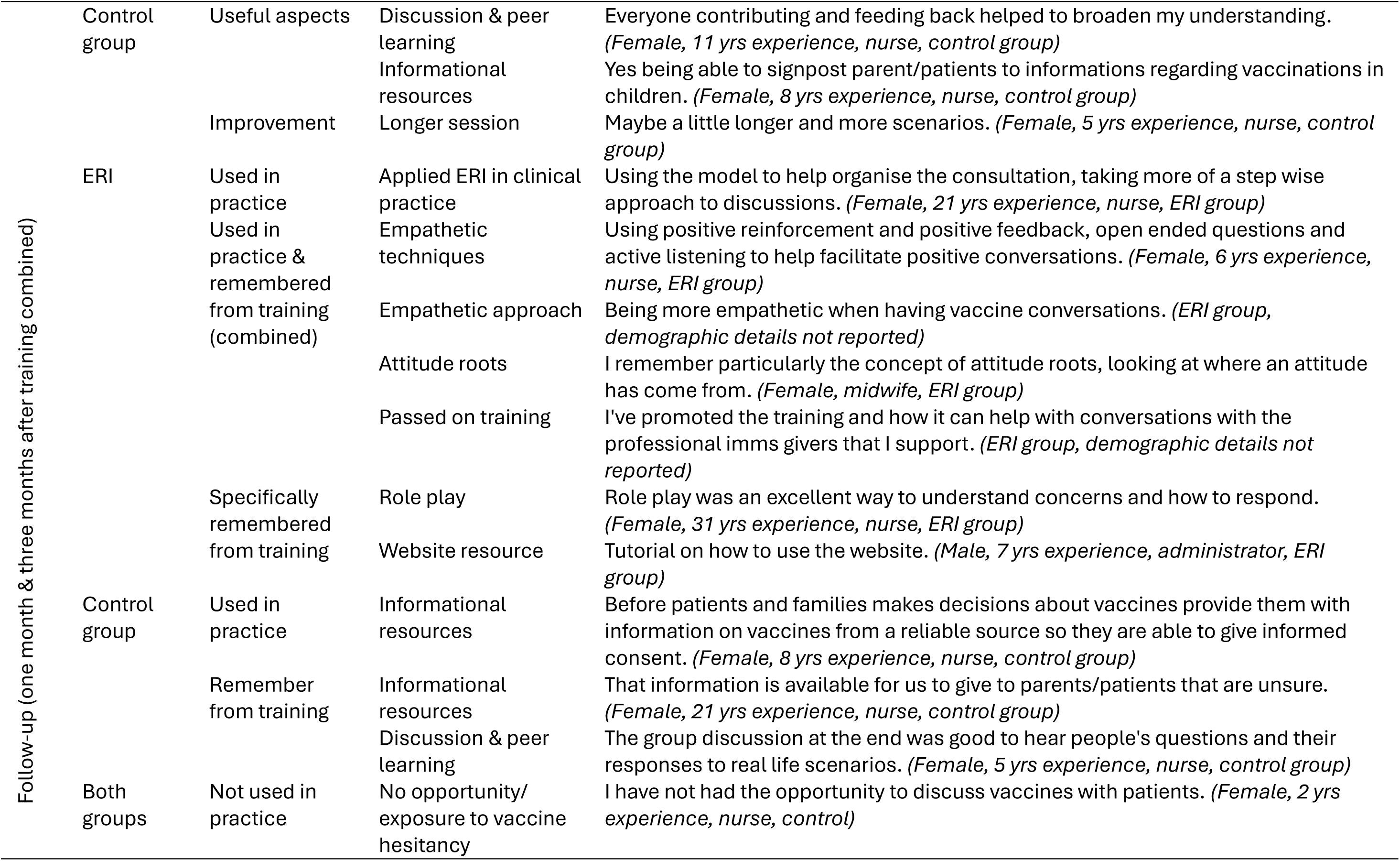
Illustrative quotations reflecting themes from qualitative data on training experience.

### Training feedback

Virtually all participants (130/131) ticked ‘yes’ to the question ‘did you find any elements of the workshop useful?’. Categories of free-text responses were similar across questions asking about usefulness of training content, improvements in understanding, intended use in practice and general feedback. As illustrated in Figure 3, a key difference between groups was that control participants described information and resources they could give their patients, while ERI participants described skills and techniques they found useful for listening to patients and guiding conversations. Thus, the control group appeared to derive confidence from having vaccine information, while the ERI group from having a way to approach a vaccine conversation.

**Figure 3.**
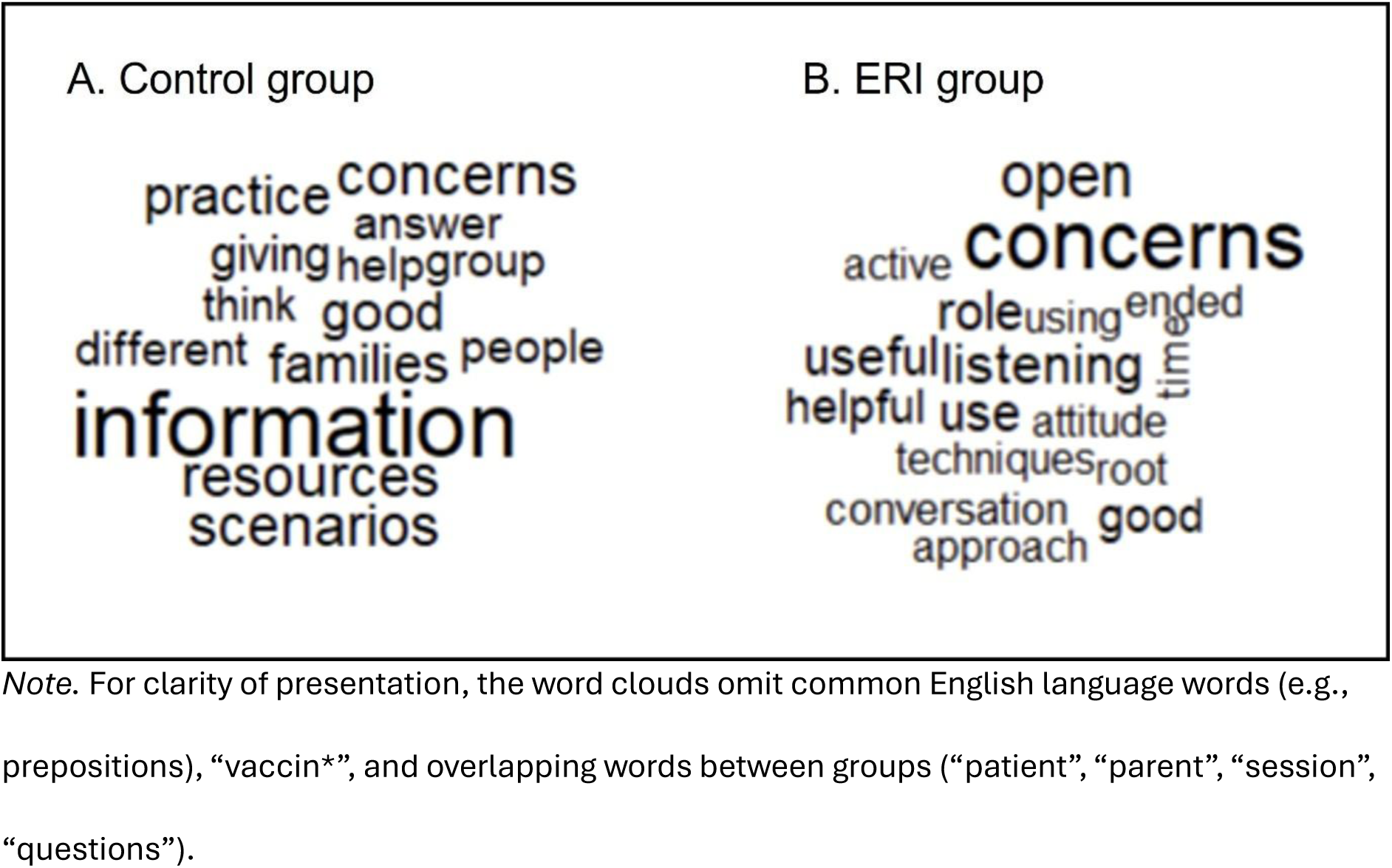
Word cloud showing the ten most frequent words in the open-ended responses from participants in the post-test and follow-up surveys.

#### ERI participants

highlighted the role play exercise as being helpful for understanding and appreciated the interactive approach and group discussion. Participants valued the framework provided by the ERI and intended to use it in practice. Many highlighted specific ERI concepts/techniques, especially affirmations and attitude roots, with some mentioning open questions and the technique of providing an alternative narrative. There were many general comments stating the training was useful, or mentioned improved understanding and confidence in how to approach vaccine conversations. A few participants mentioned the value of theory behind the ERI, and the website resource provided. Some intended to share the learning with others. General feedback was positive overall, with some appreciating the trainer’s expertise. The main suggested improvements were to have a longer session and more scenarios. Many comments suggested no improvements were necessary. A few participants wanted more facts to counter specific myths/conspiracies, or extra resources (e.g., attitudes roots handout).

#### Control group participants

mainly valued the discussion and peer learning, especially any tips on communication from experienced trainees. They also valued informational resources. They expressed confidence from being able to answer patients’ questions with facts or refer patients to information sources, which they intended to use in practice. General feedback was positive and suggestions for improvements were for a longer session and more scenarios.

Both groups commented that the role plays were helpful, though one ERI participant stated that they do not find role plays helpful, and one control participant reported being unclear on what was expected within the scenario work.

#### Practical issues

Some comments across both groups alluded to specific practical issues on the day (e.g. timing mix-up, access issue). Some (study 1) participants mentioned the session would have been better earlier in the day, and some commented on the earlier training rather than the conversation module.

### Follow up (both time points combined)

#### ERI participants

commented that they had applied the ERI framework within their clinical practice. Many mentioned the specific techniques including active listening, open questions, affirmation and taking an empathetic approach to understand patient concerns; several mentioned attitude roots in this context. A few mentioned they had passed on the concepts to others. Similar comments were given when participants were asked what they remembered from training, with the addition that many mentioned the valuable role play, and some the website resource.

#### Control participants

commented that they had used information and resources in their clinical practice. When asked what they remembered from training, they similarly mentioned vaccine information and facts. Many mentioned the discussion, peer learning and the value of understanding different perspectives and responses. Two mentioned role play.

Where participants across both groups said they had not used the skills in clinical practice, the reason given was always because they had not been exposed to vaccine hesitant people in their role in the intervening time.

### Structured observations

Observers rated module fidelity as good-excellent: all trainers covered the planned content and provided sufficient time to complete and discuss the training exercises. Observers corroborated participant feedback about course duration and timing. They noted that trainees might need more time to grasp communication skills: trainees tended to jump quickly to correcting misconceptions and providing facts when role-playing, even when explicitly instructed to focus on eliciting concerns and affirming. Observers also reflected how the module timing within the training day sometimes constrained the time available when earlier sessions in the day ran late.

## Discussion

We have shown in two studies that HCPs reported higher confidence and preparedness for vaccine conversations after short sessions about the ERI (compared to before). These improvements were comparable between the ERI and control group. However, a key difference was that HCPs in the control group gained confidence from having resources and information for patients, while HCPs in the ERI group gained confidence from having skills and techniques to guide conversations that maintain trust—which has been linked to beneficial health outcomes for patients^25^.

The value of the ERI module also appeared to be in providing structure (through the framework) to guide conversations and a novel concept (attitude roots) that helped participants to understand patients’ perspectives. The scaffolding that the ERI framework provided for role plays seemed particularly valuable in enabling trainees to practice new skills within a controlled and supportive environment—which is an important aspect of medical education^26^.

One limitation in comparing self-reported confidence gains across groups in Study 1 is that participants also spent the training day learning about immunisation theory and practice. Given how participants reported knowledge as confidence-building, earlier learning in the day may mask the contributory effect of the communication-specific module. It is promising then that Study 2’s participants (who completed only the ERI module) also reported improved communication confidence. Confidence could be a function of learning what to communicate as well as how, with the ERI module showing promise for building HCPs’ confidence and preparedness while guiding them towards more effective communication styles for vaccine hesitancy^27^.

Future work should assess what level of time commitment is required to achieve effective communication skills, as participants most frequently suggested more time to practice as a potential training improvement. Indeed, longer interventions in continuing medical education are associated with stronger effects on trainees^28^, with effective training to implement approaches such as MI often requiring several days^29^. We thus posit that the main value of a brief training module is to raise HCPs’ awareness and confidence to engage in empathetic and dialogue-based communication approaches, which further training could build on to strengthen HCPs’ skills. Due to time constraints, we were unable to assess actual conversation skills, so although we know that confidence and preparedness increased, we cannot evaluate if skills improve too.

Based on HCPs’ comments, we identified two sources of confidence for HCPs: gaining knowledge so they can give patients information, and gaining awareness of communication tools they can use to improve conversations. Future research could disentangle which of these are related to use of patient-centred communication in practice, especially given concerns that educational interventions for HCPs treat vaccine hesitancy as an information deficit problem that can be fixed through providing patients with information^10^. Nonetheless, HCPs’ self-reported confidence is an important starting point since this affects how much they recommend vaccines^12^.

## Conclusion

Short training modules on vaccine communication can improve HCPs’ confidence and preparedness to have conversations with vaccine-hesitant patients. HCPs may gain confidence from having more knowledge or from more awareness of communication tools. The ERI served as an evidence-based framework to structure training that was well-received, though longer training time is necessary to support conceptual understanding and practise skills.

## Data Availability

Materials, data, and the code to derive the reported analyses are shared on the OSF (https://osf.io/h8sv2/?view_only=5cddcd0379e445beba0333dbdc4b16c0).

## Footnotes

1 On two occasions, a randomly selected participant had to be moved to a different group to meet minimum training numbers for the control group. Excluding these participants from the data did not change any results.

2 However, only 58 completed the quantitative outcome measures in full at both pre- and post-test.

